# Study protocol for the development and internal validation of SPIRIT (Schizophrenia Prediction of Resistance to Treatment): A clinical tool for predicting risk of treatment resistance to anti-psychotics in First Episode Schizophrenia

**DOI:** 10.1101/2022.02.15.22270460

**Authors:** Saeed Farooq, Miriam Hattle, Paola Dazzan, Tom Kingstone, Olesya Ajnakina, David Shiers, Maria Antonietta Nettis, Andrew Lawrence, Richard D. Riley, Danielle A. van der Windt

**Affiliations:** School of Medicine, Keele University, Staffordshire, UK; Midlands Partnership NHS Foundation Trust, St George’s Hospital, Stafford, UK; Department of Biostatistics & Health Informatics, Institute of Psychiatry, Psychology and Neuroscience, King’s College London, University of London, London, UK; Department of Behavioural Science and Health, Institute of Epidemiology and Health Care, University College London, London, UK; Department of Psychological Medicine, Institute of Psychiatry, Psychology and Neuroscience, King’s College London, University of London, London, UK

**Keywords:** First episode schizophrenia, treatment resistant, prognostic model, decision tool, mixed methods

## Abstract

**Introduction:** Treatment Resistant Schizophrenia (TRS) is associated with significant impairment of functioning and high treatment costs. Identification of patients at high risk of TRS at their initial diagnosis may significantly improve clinical outcomes and minimize social and functional disability. We aim to develop a prognostic model for predicting the risk of TRS in patients with First Episode Schizophrenia, and to examine its potential utility and acceptability as a clinical decision tool.

**Methods and analysis:** We will use two well-characterised UK-based first episode psychosis cohorts: AESOP-10 and GAP for which data has been collected on sociodemographic and clinical characteristics. We will identify candidate predictors for the model based on current literature and stakeholder consultation. Model development will use all data, with the number of candidate predictors restricted according to available sample size and event rate. A model for predicting risk of TRS will be developed based on penalised regression, with missing data handled using multiple imputation. Internal validation will be undertaken via bootstrapping, obtaining optimism-adjusted estimates of the model’s performance. The clinical utility of the model in terms of clinically relevant risk thresholds will be evaluated using net benefit and decision curves (comparative to competing strategies). Consultation with patients and clinical stakeholders will determine potential thresholds of risk for treatment decision making. The acceptability of embedding the model as a clinical tool will be explored using focus groups with clinicians in early intervention services.

**Ethics and dissemination:** The development of the prognostic model will be based on anonymised data from existing cohorts, for which ethical approval is in place. Ethical approval has been obtained from Keele University for the qualitative focus groups within Early Intervention in Psychosis services (Ref: MH-210174). Findings will be shared through peer-review publications, conference presentations and social media. A lay summary will be published on collaborator websites.

**Strengths and limitations of this study:**

- The proposed study is the first step on the road towards the design and evaluation of a prognostic model and decision tool for the identification of treatment resistant schizophrenia. This could be informative to clinicians, patients, and their care providers in shared decision making and improvement of treatment plans.
- Individual participant data from two existing cohorts will be used to develop and internally validate the prognostic model.
- Using a mixed method design improves the ability to understand the limitations of the tool in a clinical context and create a foundation to develop it to be more effective.
- A limitation of the development of this tool is that the number of people with TRS may not be sufficiently large to consider all potential predictors for the model
- Further testing of the external validity of the prognostic model will be required

## INTRODUCTION

Treatment-Resistant Schizophrenia (TRS) is associated with the highest level of impairment of functioning amongst mental illnesses[1] and with reduced likelihood of remission and poor adherence to treatment.[2] UK estimates for the costs of TRS are not available but, considering that almost a third of patients with schizophrenia develop TRS and the total costs for the illness are estimated at about 12 billion pounds a year,[3] TRS may be responsible for at least 4 billion pounds in UK. The caregivers for TRS face significant impact on finances, career prospects, social relationships, and sense of freedom leading to frustration and hopelessness.[4]

The definition of treatment resistance and response in schizophrenia is challenging, but there is broad consensus in the literature that TRS can be defined as lack of response to treatment with at least two different antipsychotics each used for a minimum of six weeks.[5]

There is emerging evidence that TRS may be a categorically different illness subtype from the treatment-responsive schizophrenia. Lally et al.[6] examined 5-year clinical outcomes in a cohort of 246 patients with first-episode schizophrenia. Treatment resistance was seen in 34% of patients at 5-year follow-up; 70% of which did not respond to antipsychotics from the start of first treatment. In a separate cohort, Demjaha et al.[7] found that of the patients who were treatment resistant, 84% were treatment resistant from the start. A systematic review that compared baseline characteristics of patients with treatment-resistant and treatment-responsive schizophrenia reported that treatment-resistant patients showed alterations in glutamate and dopaminergic pathways, a lack of dopaminergic abnormalities, and significant decreases in grey matter compared to treatment-responsive patients,[8] which supported the hypothesis that treatment-resistant schizophrenia is a categorically different illness subtype to treatment-responsive schizophrenia.[9]

The identification of patients at high risk of TRS at the time of initial diagnosis of schizophrenia has the potential to improve clinical outcomes, prevent harm from ineffective treatment, and minimize the social and functional disability that results from prolonged psychosis,[10-11] as potentially beneficial interventions for TRS are currently not used optimally.[12] Treatment with clozapine, the only medication licensed for TRS, is often delayed between 5-10 years [13-14] and is generally prescribed to less than 10% of patients who would benefit from the drug.[12] This inadequate use of Clozapine is mainly due to mandatory blood testing required for Clozapine use, fear of serious side-effects, lack of clear guidance on identification of TRS and difficulty in selecting suitable patients.[15] Other interventions for TRS such as cognitive behavioural therapy (CBT) are used even less optimally.[16]

The early identification of those at risk of treatment resistance would minimise the delays to effective treatments for TRS, such as clozapine or CBT, thus preventing the much greater disability and morbidity associated with TRS. A number of potential predictors (prognostic factors) of TRS have been identified that can be measured at the time of diagnosis of a First Episode Schizophrenia (FES). These include poor premorbid functioning, co-morbid personality disorder, longer duration of untreated psychosis (DUP), greater severity of negative symptoms, younger age of illness onset, history of serious obstetric complications, perinatal insult and higher level of affective flattening.[17-21] Lack of response to antipsychotic medication within the first two weeks and lower remission rates at one year has also been shown to be strong predictor of subsequent poor response.[22-23] A recent systematic review identified the following predictors for TRS in FES: lower premorbid functioning; lower level of education; negative symptoms from first psychotic episode; comorbid substance use; younger age at onset; lack of early response; non-adherence to treatment; and longer duration of untreated psychosis.[24]

Prognostic models are used in healthcare to estimate (predict) the risk of future outcomes conditional on the values of multiple predictors (prognostic factors),[25] and may guide treatment decisions and prognostic stratification, for example as proposed in other long-term conditions, including musculoskeletal pain and cancer.[25] In these areas, early identification of patients likely to respond well to (or experience least harm from) certain treatments has been shown to result in improved patient outcomes and/or more efficient healthcare.[26-29]

Previous prognostic models in schizophrenia and related psychosis have been limited to a risk calculator devised for the prediction of psychosis onset[30] and a rating scale to predict the long term outcome of first episode psychosis.[31] A study that aimed to identify individuals with TRS based on genome-wide association data concluded that the use of a polygenic risk score for early identification of TRS is inadequate and not of clinical utility.[32] To address this, we propose to develop a prognostic model and develop a clinical tool for identifying patients at risk of TRS at the time of diagnosis of first episode. The model will be based on multiple predictors in combination, including clinical and sociodemographic characteristics. Then, the tool will be defined based on the model predictions and thresholds of risk that define the need for clinical action, as identified by key stakeholders. The tool will support early identification of TRS and shared decision-making regarding alternative treatment options.

This paper describes the protocol for our work, which closely follows the PROGRESS framework and guidance for prognosis research.[25,33-36]

## METHODS AND ANALYSIS

### Objectives

Our study objectives are to

i. Identify sociodemographic and clinical characteristics likely to be associated with resistance to standard (non-clozapine) antipsychotics drugs in patients with FES
ii. Develop a prognostic model for estimating an individual’s risk of treatment resistance by 5 to 10 years based on these characteristics, and to undertake internal validation of the model’s predictive performance
iii. Translate the developed model into a clinical tool and assess its potential acceptability and clinical utility among clinicians working in Early Intervention in Psychosis (EIP) services

### Study design

This will be a mixed method study. The development of the prognostic model will use individual participant data (IPD) from two existing cohort studies, with statistical methods used to produce the model equation and then to examine the model’s predictive performance in terms of calibration, discrimination and clinical utility. Consultation with patients and clinical stakeholders will be undertaken to agree the potential thresholds of risk for treatment decision making. The acceptability and perceived utility of the tool will be examined using focus group discussions with clinicians working in Early Intervention in Psychosis (EIP) services.

### Patient and public involvement and engagement

The study has been designed with input from patients and carers. Opportunities for further input to the study are embedded throughout to complement and inform key phases of the research. A patient and carer advisory group comprising 4-6 patients (people with schizophrenia/psychosis) and carers will be established; the group will be chaired by co-author DS and coordinated by Keele University’s PPIE team. Advisory group members will contribute to stakeholder engagement events to ensure the views and opinions of people with schizophrenia/psychosis and their carers are incorporated. A separate clinical expert advisory group will also be established to include local and international experts on schizophrenia to advise on identification of clinical predictors and their relevance in day-to-day clinical practice.

### Data sources

IPD from two existing cohorts (GAP and AESOP) will be used to develop and internally validate the prognostic model, based on demographic and clinical characteristics that can be routinely measured in clinical practice. Individually, these cohorts lack the sample size to provide reliable estimates of predictive performance, but in combination the two cohorts offer a rich dataset, with an extensive set of candidate predictors and large number of outcome events (cases with TRS). The cohorts are described in more detail below. Detailed information has also been collected regarding all treatments received and adherence to treatment, and important clinical outcomes, including changes in symptoms over time, hospital admission, and death.

### Start point

Participants at first contact with mental health services for psychosis.

### Identification of candidate predictors

We will use an iterative approach to identifying candidate predictors. This will consist of following:

I. **We will identify a list of candidate predictors of treatment resistance from available literature**,[22-23,37-39] including relevant systematic reviews.[24,40] Additional focused searches (Medline, EMBASE, PsychINFO) will be carried out to identify additional evidence for candidate predictors
II. **Consultation with our clinical expert and patient advisory group: A group of experts in** experts in schizophrenia research is identified and will be consulted. The predictors identified from the literature search will be shared with this group and their views will be sought about the usefulness and validity of these factors in predicting TRS, and whether they can suggest any other potential predictors

The results of this process will inform the selection of candidate predictors to be considered in the development of the proposed prediction model, restricting to the subset that are available in both data sources for the model development. Additional predictors assumed to be important by our experts and patient advisors but not available in the two data sources can be considered for future updates or improvements of the model, if needed, as new data become available.

Once the set of candidate predictors is established, we will then further consult our clinical expert and patient advisory groups to decide on those to prioritise for inclusion in the prognostic model development (as the total will need to consider sample size restrictions in order to reduce potential for overfitting). This process will use face-to-face (patient advisory group) and videoconference (clinical expert advisory group) meetings, in line with current pandemic restrictions. These meetings will be facilitated by an experienced qualitative researcher (TK), clinical member of the study team (SF) and – for the patient advisory group – by the PPIE team member (DS). The advisory groups will be presented with a list of all candidate predictors available in the individual participant data (IPD) from two datasets (see below), along with a description of how the predictors have been measured and a brief summary of the existing evidence for their association with resistance to anti-psychotic treatment. Discussions will be audio-recorded, and written reports drafted summarising feedback and advice that ranks the predictors by their perceived prognostic importance. The agreed list of candidate predictors will then be taken forward for model development (objective ii).

### Treatment resistance definition

The definition of treatment resistant schizophrenia varies in different studies but there is broad consensus in the literature that TRS can be defined as lack of response to treatment with at least two different antipsychotics, each used for a minimum of six weeks.[5] This requires treatment with two antipsychotics using a therapeutic dose of each drug in a sequential manner, which is rare in clinical practice for number of reasons. There is usually a delay of up to five years in starting the gold standard treatment for TRS, Clozapine.[13-14]

Therefore, in the two data sources for our model development, we define the TRS cases as those who did not respond to 2 consecutive antipsychotic medication of adequate dose and for an adequate duration and/or the documented reason for switching antipsychotic medication was due to a lack of therapeutic response.[6-7,41-43] An adequate daily dose of antipsychotic medication was defined according to a daily dose of ≥400mg chlorpromazine equivalence.[44] We only included as TRS cases those patients who failed to respond and not those who were intolerant of antipsychotic medications or those who self-discontinued antipsychotic medication.[6-7,41-43] Non-response here was measured as not experiencing improvement in symptoms and social functioning in duration of 6 months during the follow-up period.[45]

### Study sample

#### Genetics and Psychosis (GAP) study

In the GAP cohort, the study sample comprised of participants aged 18-65 meeting criteria for first episode psychosis disorders (International Classification of Diseases, 10th-Revision (ICD-10) diagnoses: F20.0, F25.0, F28.0, F29.0), [46] validated by administration of the Schedules for Clinical Assessment in Neuropsychiatry. [47] All cases had been admitted to psychiatric inpatient units or seen by community-based mental health teams within the South London and Maudsley (SLaM) NHS Foundation Trust between December 2005 and October 2010.[48] The study exclusion criteria were evidence of 1) psychotic symptoms precipitated by an organic cause; 2) evidence of transient psychotic symptoms resulting from acute intoxication as defined by ICD-10; 3) moderate or severe learning disabilities as defined by ICD-10; or 4) head injury causing clinically significant loss of consciousness. Approximately 5 years after first contact with mental health services for psychosis, all patients with baseline diagnosis of schizophrenia spectrum disorders and who had given consent for follow up and for their clinical records to be accessed were followed-up. The follow-up data were extracted retrospectively using the electronic psychiatric clinical records (EPCRs). The EPCRs are the primary clinical records keeping system within the SLaM Trust that allows to search all clinical information, including correspondence, discharge letters and events, recorded throughout patients’ journeys through the SLaM Trust services. All deaths and emigrations up to and including those that occurred during the final year of follow up were identified by a case-tracing procedure with the Office for National Statistics (ONS) for England and Wales and the General Register Office (GRO) for Scotland. Overall, 246 out of 283 participants were successfully followed-up (86.9%); of these sufficient, information on treatment received was available for 239 cases. Eighty (33.5%) of the cases met criteria for TR and 159 (66.5%) were non-TR.

#### AESOP-10

Aetiology and Ethnicity in Schizophrenia and Other Psychoses (AESOP-10) is a 10-year longitudinal, population-based study of incident cases of psychosis from defined catchment areas.[7] At baseline, all patients aged 16–64 years who presented with first episode psychosis over a 2-year period in centres in southeast London and Nottingham (UK) were invited to take part at approximately 10 years post-inclusion. Out of a total of 557 recruited at baseline 434 participants provided follow-up data (77.8%). For 286 participants whom there was complete information on medication, adherence to treatment and symptom ratings over the 10-year follow-up period. Psychopathology was assessed using the Schedules for Clinical Assessment in Neuropsychiatry.[47] A diagnosis of schizophrenia was made according to International Classification of Diseases (ICD)-10 diagnostic criteria for research during clinical consensus meetings. Ethical approvals for both cohort studies were obtained from local research ethics committees.

Following variables are recorded in both datasets at baseline: Age at first contact with mental health services, gender, ethnicity, baseline diagnosis (IC-10 and DSM), duration of untreated psychosis (DUP), IQ, educational level, employment status, family history of psychosis, substance abuse, alcohol use, symptoms dimensions, childhood adversity, mode of onset, living arrangements and living alone, being single or separated. To ensure minimal exposure to antipsychotic medications in patients, all assessments were obtained within a 3-month period following the first contact with psychiatric services.

### Outcome measures

For this study, the primary outcome measure will be the occurrence of treatment resistance schizophrenia during the follow up period, as defined above. We will examine the prediction of treatment resistance by 5 to 10 years. As the follow-up period is 5 years in GAP and 10 years in AESOP-10, and outcome status is unknown by 5 years (AESOP-10) or 10 years (GAP), in order to utilise both datasets for model development we make the assumption that risks by 5 and 10 years are similar. This will need to be checked in future external validation studies.

### Sample size for model development

The combined dataset includes 532 (286 AESOP + 246 GAP) participants with complete follow-up data, and sufficient information on drug treatment and symptom severity to define treatment resistance Across both cohorts a total of 155 (29%) participants were identified with TRS based on previous studies.[7] The number of predictor parameters will be restricted to adhere to this sample size and outcome events (not considering additional statistical power that may arise following inclusion of those with missing TRS values based on multiple imputation). Based on the evidence identified in our literature reviews, we will extract the potential Cox-Snell R-squared for models in this field, and use it to inform the maximum number of candidate predictor parameters that can be considered for inclusion in the model whilst minimising overfitting.[49] For example, if the Cox-Snell R-squared may be about 0.22 for the new model, then up to 15 potential predictor coefficients could be considered, as 15 is the maximum required given the 532 sample size and 29% outcome proportion.[49-50] The sample size also ensures the overall outcome proportion will be estimated precisely, with upper and lower values of the 95% confidence interval within 0.04 of the outcome proportion of 0.29.[50]

### Statistical analysis

#### Model development and internal validation

Development and internal validation of the prognostic model will be guided by the PROGRESS framework,[33-34,50] and will be reported adhering to the TRIPOD guidelines.[51]

First, the extent and distribution of missing data for predictors, treatment characteristics, and clinical outcomes will be described. For each cohort, baseline characteristic of participants with complete follow-up data will be described and compared to those who were lost to follow-up in order to assess the risk of attrition bias. Imputation of missing data will be handled separately for each cohort, prior to combining the data for predictive modelling, to retain any potential between-cohort heterogeneity. Imputation of missing data (both outcomes and predictors) will be handled using multiple imputation and Rubin’s rules, under a missing at random assumption, including outcome and candidate predictors in the imputation model alongside a broader set of auxiliary variables available in each cohort (to improve the missing at random assumption).[52-53]

Next, a model for predicting risk of TRS will be developed based on penalised multivariable regression, considering all candidate predictors emerging from the stakeholder meetings with clinical expert and patient advisory groups, as described above and adhering to the sample size calculation. As there is no censoring in the datasets, we will use logistic regression for occurrence of treatment resistance over the follow-up period (5 to 10 years).

All predictors chosen for inclusion (adhering to the sample size criteria) will be forced to be included, given the consensus process to identify them as candidate predictors, and to remove the potential for instability caused by variable selection methods. Continuous variables will not be categorised and potential non-linear effects examined using splines or fractional polynomials.

Apparent performance will be summarised in terms of calibration (calibration plots with smoothed calibration curves; calibration slope and calibration-in-the-large) and discrimination (C-statistic, area under the curve). Then, internal validation will be undertaken using bootstrapping of the entire development dataset (accounting for the clustering of participants within studies), to estimate optimism in model performance, and to then derived optimism-adjusted estimates of predictive performance for calibration (e.g. calibration-in-the-large, calibration slope) and discrimination (C-statistic) of predicted risks. Finally, using the optimism-adjusted calibration slope as a global shrinkage factor, we will shrink (penalise) predictor effects for overfitting. Following this the intercept term will be re-estimated to ensure calibration-in-the-large.

Between-cohort heterogeneity in baseline risk, predictor effects and the model’s predictive performance will be examined in both apparent and interval validation approaches.

### Deciding thresholds for treatment decision making based on the prognostic model

The prediction model will generate the probability (risk) of TRS for individual patients, which in itself (following external validation of predictive performance) will be of future importance to clinicians, patients, and their care providers, as it will provide information on prognosis, and can inform discussions regarding different treatment options. However, our aim is to use the prognostic model not just for individual risk prediction, but also as a clinical decision tool to guide decisions regarding alternative treatment options (e.g. start Clozapine or continue other antipsychotics) in those at high risk of treatment resistance. This translation will require the identification and agreement of thresholds of predicted risk above which a change in treatment (i.e. transfer to clozapine) is warranted and worthwhile, considering the potential benefits, harms (adverse effects) and costs of treatment. For deciding on such thresholds we will use decision analysis and stakeholder involvement (see below).

### Net benefit and decision analysis

Evidence regarding the effects and harms of clozapine therapy for patients with schizophrenia will be extracted from existing (Cochrane) systematic review/meta-analysis evidence.[11,54-56] Assuming intervention effects are constant across individuals on a relative scale, those at high risk of TRS are more likely to benefit from treatment in terms of absolute risk reduction than those with low predicted risk of TRS. The thresholds at which clozapine prescription is indicated will need to be determined, based on an assessment of the balance between expected benefits and risks, in consultation with our clinical and patient advisory groups. Using lower thresholds for predicted probabilities will increase sensitivity and miss fewer patients likely to be resistant to first-line antipsychotic treatment, but inevitably means that a larger number of patients without treatment resistant schizophrenia (TRS) would unnecessarily require treatment with clozapine, and vice versa.

After model development is completed, optimism-adjusted decision curves and net benefit will be calculated to quantify clinical utility across the range of risk thresholds deemed relevant by the advisory group.[57] The scenarios used in this analysis will be based on real-life case descriptions, developed together with our patient and expert advisors (see below). The decision analysis therefore will be based on actual clinical and lived experience of TRS, and include the social and clinical implications of clozapine use for young individuals when discussing the balance of expected benefits and harms of clozapine therapy compared to first-line antipsychotics or other treatment options. Comparison of net benefit and decision curves for the new tool will be made with other strategies of ‘treat all’ and ‘treat none’.

### Consensus exercise with clinical and patient and carer advisors

A workshop will be organised with members of our patient and clinical expert advisory groups, and clinicians from local psychiatric intervention services including about 15 participants. The final prognostic model and its predictive performance will be presented along with results of the decision model. We will use graphics that are accessible to both clinicians and patient representative, in order to illustrate the expected benefits and harms of clozapine at different levels of predicted risk of treatment resistance. We will use an adapted Nominal Group Technique (NGT) as a consensus building exercise to agree optimal thresholds for clozapine prescribing. The NGT is a group process involving problem identification, solution generation, and decision-making through ranking or voting that is time-efficient and involves low-burden for participants.[58-59] It allows a rapid process of decision-making, while taking every workshop participant’s opinion into account.

The following issues will be discussed: (i) predictive performance of the model; (ii) profiles of subgroups of patients with schizophrenia at varying levels of predicted TRS risk; (iii) effect estimates of clozapine at varying levels of predicted TRS risk; (iv) balance of benefits, risks, and costs of clozapine treatment at varying levels of predicted TRS risk, before achieving consensus regarding proposed thresholds for clozapine prescribing based on the prognostic model. The meetings will be facilitated by an experienced researcher, clinical member of the study team, and PPIE team member. Meetings will be audio-recorded and transcribed to ensure the decision-making process is captured, including the different views prior to agreement regarding each factor.

### Evaluating acceptability and clinical utility of clinical decision tool

The acceptability and anticipated clinical usefulness of the clinical tool will be assessed using focus groups with clinicians in Early Intervention in Psychosis (EIP) services. Focus groups provide a useful means of facilitating discussion between participants to explore specific issues in health research.[60]

Two focus groups will be conducted each consisting of 6-10 participants. Potential participants will be invited based on the following criteria: (i) are a qualified psychiatrist; (ii) work in early intervention services or in-patients with responsibility for admission of early psychosis patients; and (iii) are involved in the diagnosis and management of schizophrenia. Potential participants will be identified through local EIP services in Stafford and London. Potential participants will be sent an information sheet and ‘consent to further contact’ form to complete and return to the research team. A date will then be arranged at a convenient location for the participants (e.g. NHS site).

Informed consent will be obtained at the start of the focus group, which will be digitally recorded. Demographic information will be collected to support description of the participants, including: sociodemographic characteristics, number of years’ experience as a psychiatrist, location of practice and number of patients with schizophrenia. An experienced qualitative researcher will facilitate the focus group using a semi-structured topic guide. Participants will be asked for their views on TRS and their clinical approach to managing TRS. Participants will also be asked to give their views on two case vignettes that describe the characteristics of patients at low and high risk of predicted TRS, including the results of the clinical prediction tool and linked treatment recommendations (based on the results from the workshop with our patient and expert advisory groups). The two vignettes will be sent to the participants prior to the focus group. The following topics will be explored through discussion among participants: content of the tool and wording of items; usefulness of the clinical scale in clinical practice to assess TRS; usefulness of the tool to guide shared decision making regarding treatment; acceptability to service users; methods of implementing the tool in clinical practice (paper or electronic, linkage to medical records); need for instructions.

### Qualitative analysis and co-design of the clinical decision tool

Digital recordings from the focus groups will be transcribed verbatim for analysis. Thematic analysis will be conducted to identify and explore important themes from the data.[61] The qualitative researcher will lead analysis with clinical and non-clinical members of the research team, and input from the PPIE study team member, contributing to the interpretation of data through analysis meetings; this mixed team collaborative approach helps to support the trustworthiness of the finding.[62]

The qualitative findings will then be discussed during a workshop with members of our patient and clinical expert advisory groups (n≈10 participants), and a final version of the tool, including optimal ways for dissemination and implementation will be agreed. A proposal for further validation and impact analysis will be developed, aiming to investigate the impact of using the tool on patient outcomes and costs of care.

The above process will result in an agreed clinical tool for assessing treatment resistance in first-episode schizophrenia that may be used to support decisions regarding transition to treatment with clozapine for those at high risk; (ii) information on potential acceptability and clinical utility of the tool in terms of net benefit (over other strategies such as treat all and treat none); (iii) suggestions for dissemination and implementation in routine clinical practice; (iv) recommendations for future research aiming towards further validation of the tool and investigation of clinical and cost-effectiveness of using the tool in the management of first-episode schizophrenia.

## ETHICS AND DISSEMINATION

### Obtaining approvals

The development of the prognostic model will be based on anonymised data from the existing AESOP and GAP cohorts, for which ethical approval is in place. Formal data request procedures have been followed to enable access of anonymised data by the analysist team at Keele University. Ethical approval has been obtained from Keele University for the qualitative focus groups with Early Intervention in Psychosis services (Ref: MH-210174).

### Dissemination and impact

All outputs will be produced in consultation with service users. The proposed research will include two outputs: (i) a prediction model designed to estimate the probability of TRS using information available at the time of diagnosis of schizophrenia, and (ii) a clinical decision tool, i.e. the prediction model plus an agreed set of thresholds to offer clozapine rather than first-line antipsychotics, including balance of expected costs, benefits and harms of clozapine therapy at these thresholds. The results of the study will be reported in peer reviewed scientific journals, presented at major national and international conferences, and loaded on our websites (Keele and King’s College). Lay summaries will be prepared in collaboration with our PPIE representatives (patient advisory groups) and disseminated through our website, and social media, via our dedicated and widely followed Twitter and Facebook feeds. The team have strong links with professional organisations and networks e.g. The Royal College of Psychiatrists and IEPA, an international organisation of early intervention in psychosis. These will be used for further dissemination of our findings.

## DISCUSSION

The clinical tool to predict treatment resistance in schizophrenia will be a significant advance in management of schizophrenia and developing prognosis research in schizophrenia. The proposed study is the first step on the road towards the design and evaluation of a prognostic model and decision tool for the identification and management of treatment resistant schizophrenia. The model will allow the probability (risk) of TRS to be estimated for an individual patient, which in itself could be informative to clinicians, patients, and their care providers in shared decision making. In particular, by providing information about likely prognosis with anti-psychotics for treatment after diagnosis with a FRS, and can inform discussions regarding different or additional treatment options.

WHO framework of prevention of mental disorders[63] proposes primary prevention seeks to prevent the onset (incidence) of a disorder or illness while secondary prevention seeks to lower the rate of established cases of the disorder or illness in the population. Both are prohibitively difficult in psychiatric disorders in view of the limited predictive strength of all known risk factors for mental disorders.[64] For example, testing positive for high risk for psychosis has been found to be associated with a 6% lifetime risk of actually developing psychosis.[65] This means that 94% of those who score positive will not develop psychosis in their lifetime. Preventive interventions for such a small proportion of at risk population will be ethical and economically challenging.[64] Tertiary prevention aims to reduce disability and enhance rehabilitation in long term and chronic condition.[63] Successful development of SPIRIT will result in early identification of people with schizophrenia who are not likely to respond to standard antipsychotics. This can potentially prevent a debilitating and chronic psychosis in the form of TRS and present an opportunity for tertiary prevention.

Further development and testing of the tool will depend on the actual results of the project. If the results of the proposed study show the model has good predictive performance, we propose testing of the external validity of the prognostic model (in regards to both 5 and 10 year outcomes) and confirmation of decision thresholds.

After further validation, the prognostic model and linked decision tool could potentially identify patients who would benefit from interventions for TRS at the earliest possible stage, thus improving the quality of health services with potential reduction in overall costs. This will help to reduce symptoms, overall morbidity and improve the quality of life. Early identification and treatment for TRS can potentially lead to other wide ranging positive outcomes, including increased awareness, improvements in relationships and greater employment.

The involvement of service users in the study will improve the participants’ understanding of research in TRS and raise general public awareness which will guide people to seek early clinical help for their mental health problems.

If study aims are met, the research will inform the methodology for prognosis research in early psychosis and Psychiatry in general. It will also contribute to the development of research aiming to target effective treatment at the earliest possible stage of schizophrenia, possibly triggering future research such as a full impact analysis study (e.g. cluster-RCT, or large comparative observational study) to investigate the clinical and cost-effectiveness of using the decision tool on patient outcomes and costs of care.

## Data Availability

This is inapplicable as this is a protocol paper and data has not yet been produced

## Author contributions

SF, PD, OA, DS, RDR, DvdW and TK were involved in the original conception and design of the research. All authors SF, MH, PD, TK, OA, DS, MAN, RDR, DvdW have contributed to the drafting of this manuscript.

## Funding statement

This study is supported by NIHR Research for Patient Benefit Programme grant number [NIHR 200510]. OA is funded by an NIHR Post-Doctoral Fellowship (PDF-2018-11-ST2-020).

## Notes

### Competing Interest Statement

The authors have declared no competing interest.

### Funding Statement

This Study was funded by the NIHR

### Author Declarations

The Health research authority - 21/HRA/3472 gave ethical approval for this work Keele University FMHS Faculty Research Ethics Committee - MH-210174 - gave ethical approval for this work

